# Analysis of Differentially expressed genes and Molecular pathways in Peanut Allergy Induced Dynamic Changes involved in Peanut-Specific Immune Responses: A Systematic and Bioinformatics Approach

**DOI:** 10.1101/2024.05.12.24307235

**Authors:** Glory Simon Parmar, Manisha Gulati, Md Khoshnade Rabby, Ramchander Merugu, Satabdi Mohanty, Umme Kulsum, Dhanshree Gajanan Pujari, Swapnja Rohit More, Om Prakash, Priyanka Shrikant Bhore, Deepshikha Satish

**Author notes:** is corresponding Author. Authors have contributed equally (Glory Simon Parmar, Manisha Gulati, Md Khoshnade Rabby, Ramchander Merugu, Satabdi Mohanty, Umme Kulsum).

## Abstract

Peanut allergy poses a significant global health concern, triggering hypersensitivity reactions upon exposure to peanuts. Understanding the molecular mechanisms governing peanut-specific immune responses is imperative for developing effective therapeutic approaches. This study aimed to investigate differentially expressed genes (DEGs) and associated molecular pathways implicated in peanut allergy-induced immune responses. Employing a systematic and bioinformatics-driven approach, we analyzed gene expression profiles from peanut-allergic individuals and healthy controls using RNAseq Next-Generation Sequencing (NGS) data. Enrichment analysis of DEGs revealed their involvement in various inflammatory conditions, including autoinflammatory, allergic, and respiratory disorders. Additionally, pathway enrichment analysis highlighted perturbed molecular pathways such as Asthma, IL-17 signaling pathway, and Inflammatory bowel diseases, underscoring their role in modulating peanut-specific immune responses. Protein-protein interaction network analysis identified central regulatory hubs, elucidating the intricate molecular interplay underlying the immune response to peanut allergens. Overall, our findings offer comprehensive insights into the molecular landscape of peanut allergy, delineating key DEGs and pathways associated with peanut-specific immune responses. This systematic and bioinformatics-driven approach enhances our understanding of allergic reactions to peanuts, providing potential targets for the development of novel therapeutic interventions and diagnostic biomarkers for peanut allergy management.

## 1. INTRODUCTION

Peanut allergy presents a significant global health challenge, particularly prominent in Western societies, where its incidence has surged over recent decades[1]. Marked by heightened sensitivity to peanut proteins, this allergy carries a substantial risk of severe, potentially life-threatening reactions, underscoring the importance of unraveling the molecular mechanisms governing immune reactions to peanuts. The pathogenesis of peanut allergy involves a multifaceted interplay of genetic predisposition, environmental influences, and immune system dysregulation. While the precise mechanisms remain partly elusive, recent strides in molecular biology and bioinformatics have furnished indispensable tools for delving into the intricate molecular pathways and identifying pivotal genes implicated in the immune responses specific to peanuts. This analysis lays the foundation for comprehending the development of allergies stemming from peanuts.[2]

The analysis of RNA-seq experiments requires precise and efficient software capable of managing the substantial sequencing volumes typically generated[3]. This process involves four primary tasks: aligning reads to the genome, assembling alignments into full-length transcripts, quantifying gene and transcript expression levels, and calculating expression differences across experimental conditions. STAR efficiently aligns RNA-seq reads to the genome, discovering transcript splice sites while running much faster[4]. Cuffdiff, built upon Cufflinks, rigorously identifies differentially expressed transcripts between samples using statistical methods[5]. This paper presents a protocol for utilizing these tools in RNA-seq data analysis, all of which are thoroughly documented and actively maintained by developers.

This study focuses on analyzing differentially expressed genes (DEGs) and molecular pathways associated with peanut allergy-induced changes in immune responses. Through transcriptomic analysis of peanut-allergic individuals compared to non-allergic controls, we aim to identify genes significantly altered in response to peanut allergen exposure. These DEGs hold potential as biomarkers and therapeutic targets for peanut allergy. We will annotate the biological functions of identified DEGs and assess their enrichment in specific molecular pathways, unraveling the biological processes driving peanut-specific immune responses and allergic reactions. Integrating transcriptomic, proteomic, and metabolomic data will provide a comprehensive understanding of molecular networks governing peanut allergy, revealing gene-gene interactions and regulatory mechanisms contributing to its pathogenesis[6].

Candidate DEGs and molecular pathways identified through bioinformatics analysis will undergo experimental validation using in vitro and in vivo models of peanut allergy, providing functional insights into their role in modulating immune responses. Computational analysis enables the use of gene expression differentials for functional annotation, pathway analysis, and molecular integration. Our research aims to elucidate the molecular mechanisms underlying peanut allergy, informing the development of targeted therapeutic interventions and diagnostic strategies. By combining sequencing technologies with rigorous analysis, we aim to uncover genetic intricacies within peanut samples, advancing personalized medicine and biological research[7].

## 2. MATERIALS AND METHODS

### 2.1. Data Acquisition and Processing

The methodology initiated with the acquisition of raw RNA-seq reads sourced from the Sequence Read Archive (SRA) database (https://www.ncbi.nlm.nih.gov/sra), specifically utilizing the identifier SRX14128943 for investigating peanut allergy. Datasets pertinent to various experiments were gathered by accessing gene expression data stored within the database. The retrieval process involved querying the SRA database using keywords such as “Food allergen,” “RNAseq,” and “Homo sapiens” to identify datasets featuring relevant gene expression profiles. The dataset under investigation, SRX14128943, comprised twenty-five samples, encompassing seven and nineteen peanut allergens, as delineated in Table 1. Subsequently, gene expression data were extracted from the European Nucleotide Archive (ENA) database (https://www.ebi.ac.uk/ena/browser/).

**Table 1:** List of selected individuals from the SRA Database, allergic to peanuts, with a control group of healthy individuals.

| Sr No | Sample ID | Sample Type |
| --- | --- | --- |
| 1 | SRR17972672 | Control |
| 2 | SRR17972673 | Control |
| 3 | SRR17972674 | Control |
| 4 | SRR17972675 | Control |
| 5 | SRR17972676 | Control |
| 6 | SRR17972677 | Peanut Allergic |
| 7 | SRR17972678 | Peanut Allergic |
| 8 | SRR17972679 | Peanut Allergic |
| 9 | SRR17972680 | Peanut Allergic |
| 10 | SRR17972681 | Peanut Allergic |
| 11 | SRR17972682 | Peanut Allergic |
| 12 | SRR17972683 | Peanut Allergic |
| 13 | SRR17972684 | Peanut Allergic |
| 14 | SRR17972685 | Peanut Allergic |
| 15 | SRR17972686 | Peanut Allergic |
| 16 | SRR17972687 | Peanut Allergic |
| 17 | SRR17972688 | Peanut Allergic |
| 18 | SRR17972689 | Peanut Allergic |
| 19 | SRR17972690 | Peanut Allergic |
| 20 | SRR17972691 | Peanut Allergic |
| 21 | SRR17972692 | Peanut Allergic |
| 22 | SRR17972693 | Peanut Allergic |
| 23 | SRR17972694 | Peanut Allergic |
| 24 | SRR17972695 | Peanut Allergic |
| 25 | SRR17972696 | Peanut Allergic |

### 2.2. Data Preprocessing and Identification of DEGs

Utilizing advanced Next-Generation Sequencing (NGS) technology, the initial dataset underwent rigorous quality assessment using FastQC (v0.11.9)[8], followed by adapter trimming via Trimmomatic0.39. Subsequently, reads from each sample were aligned to the reference genome using STAR, yielding sorted BAM files[4]. These alignments were then subjected to normalization and abundance estimation using Cufflinks (v2.2.1), enabling the quantification of gene and isoform expression levels[5]. Cuffmerge was employed to assemble genes for each dataset individually. Post-assembly, the complete set of assemblies was processed through Cuffdiff to compute differential gene expression, with resultant p-values and false discovery rates (FDR/q-values) determining significant differential expression between peanut allergens and controls[9].

We followed the stringent criteria, requiring |log(fold change)| > 1 and p < 0.05 to identify significant DEGs from the dataset. Upregulated DEGs were defined by logFC ≥ 1, while downregulated DEGs were identified by logFC ≤ -1. Subsequently, RStudio (v4.3.2) and the ggplot2 package were utilized to generate a volcano plot visualizing the significant DEGs. Identified DEGs were extracted from the dataset for further analysis.

To visualize gene expression patterns, a heatmap was constructed using the ComplexHeatmap library. This protocol is accessible without specialized programming skills but assumes familiarity with the Unix command-line interface and basic R scripting[10]. Users should feel comfortable executing commands in the Unix environment and editing text files. The study workflow is illustrated in Fig 1.

**Fig 1.**
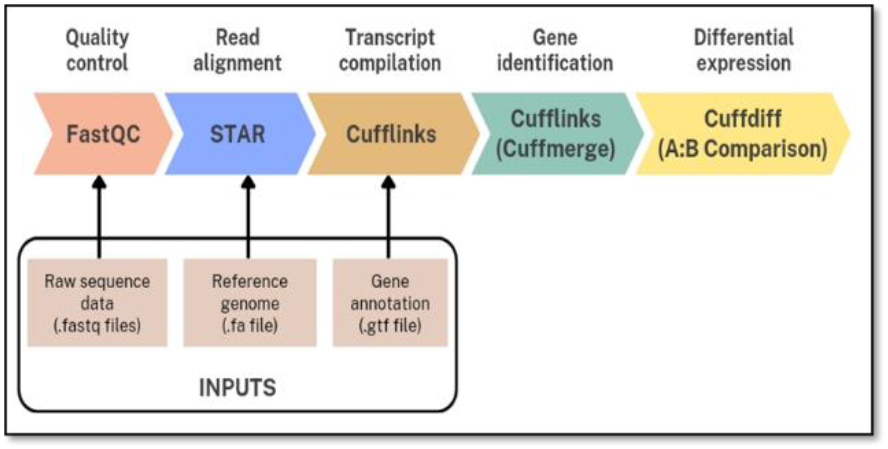
Shows RNA-Seq pipeline begins with quality control, which used FastQC to check the sequencing data’s quality, assuring reliable downstream analysis. The readings were then aligned to a reference genome using STAR, which allows for more accurate mapping of RNA transcripts. Cufflinks are then used to assemble transcripts, allowing potential transcripts to be reconstructed from matched reads. Cuffmerge is used to identify genes by aggregating transcript annotations into a single set for further analysis. Finally, Cuffdiff is used for differential gene expression analysis, which allows the identification of genes that are differentially expressed across experimental conditions, providing insights into biological mechanisms.

### 2.3. Functional Enrichment Analysis

Functional annotation and analysis of KEGG pathway enrichment were conducted using the web-based DAVID v6.8 tool, a reliable resource for comprehensive functional evaluation of high-throughput gene expression data[11]. The outcomes from DAVID were subsequently imported into R Studio for visualization using ggplot2. Additionally, ShinyGO (v 0.80) was employed to visualize functional and pathway enrichment of the top 10 Differentially Expressed Genes (DEGs). ShinyGO facilitates the integration of expression data with functional assessment results, enabling a deeper understanding of molecular, biological, and cellular functions, as well as pathway enrichment analysis[12].

### 2.4. Protein-Protein Interaction (PPI) Network Construction and Module Analysis

The protein-protein interaction (PPI) network was constructed using the STRING web-based tool (v12.0) to elucidate relationships among the DEGs identified in the datasets[13]. A medium confidence interaction score cutoff of ≥0.4 was applied to filter out inconsistent PPI interactions from the dataset, resulting in the establishment of a robust PPI network. Subsequently, the outcomes from the STRING tool were integrated into Cytoscape software (v3.8.2) to visualize the PPI interactions among statistically significant DEGs. Hub genes within the PPI network were identified by sorting based on the degree of interaction. For enhanced visualization, the yFiles organic layout was applied to represent the network structure effectively.

## 3. RESULTS

### 3.1. Identification of Differentially Expressed Genes (DEGs) and Data Visualization

The gene expression profiles of nineteen peanut allergen samples and seven control groups (CD154+ CD4 T-cells) were acquired from the SRA database (SRX14128943). Utilizing the Illumina HiSeq 2500 platform, DEGs between control samples and peanut allergens were identified employing the cuffdiff tool. DEGs were determined based on the criteria of | log2FC| ≥ 1.0 and p-values ≤ 0.05. A total of 444 genes were identified as biologically significant. Subsequently, a volcano plot was constructed using R studio and the ggplot2 package, visually representing the DEGs. In the plot (Fig 2), upregulated significant genes are depicted as red dots (p-value ≤ 0.05, logFC ≥ 1), while downregulated significant genes are shown as green dots (p-value ≤ 0.05, logFC ≤ -1). Furthermore, the top 20 DEGs were subjected to a heat map analysis using the ComplexHeatmap package, revealing the expression levels of these genes (Fig 3).

**Fig 2:**
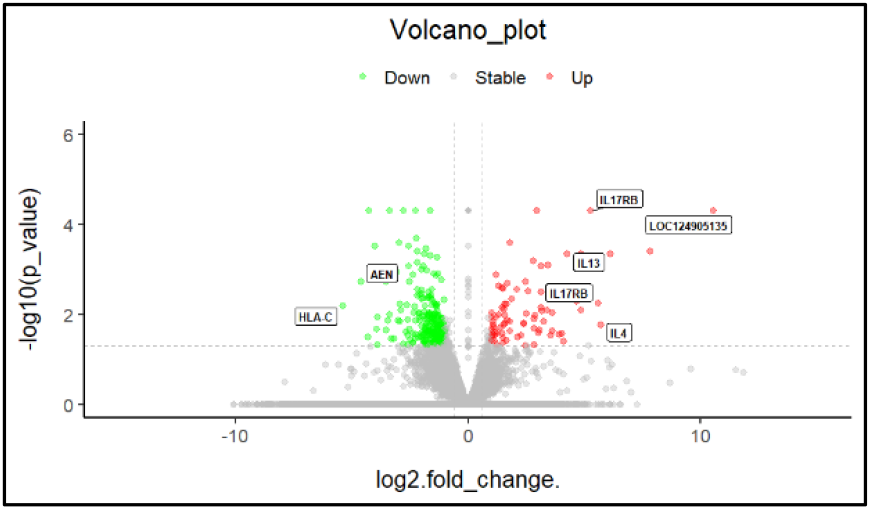
A volcano plot is a popular graphical depiction for differential gene expression studies. It shows fold change on the x-axis and statistical significance (typically written as -log10(p-value) on the y-axis. Genes with significant changes in expression are shown as points above a specific threshold, which are often highlighted in the plot, generating a volcano-like shape with the most significant genes at the top.

**Fig 3:**
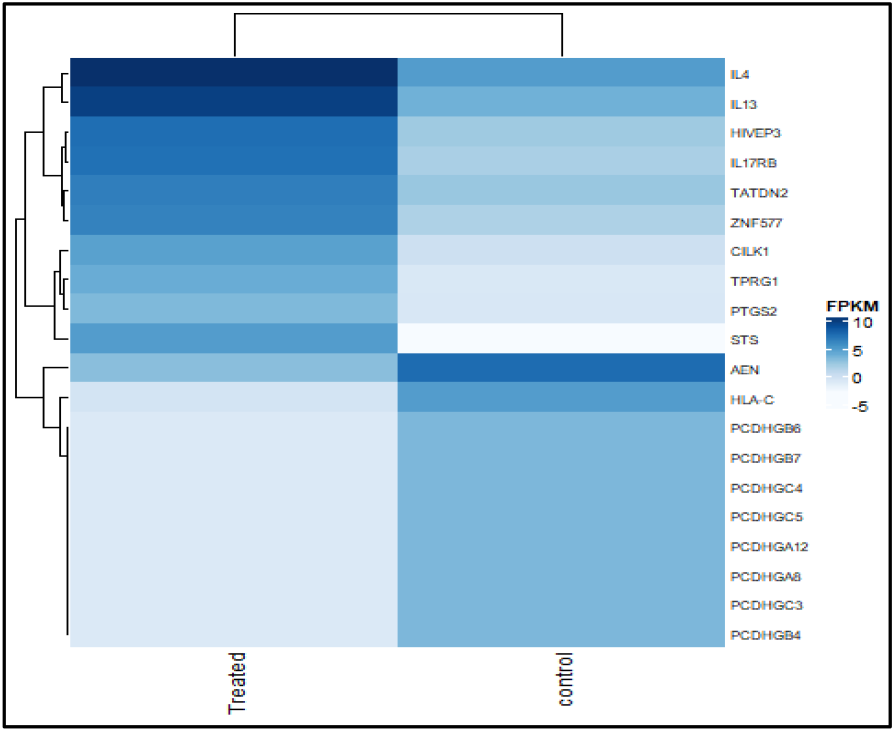
A heat map of gene expression is a visual representation of gene expression levels across various samples or conditions. Normally, rows indicate genes, columns represent samples, and colors represent expression levels, with brighter colors suggesting higher expression and darker shades indicating lower expression. Heat maps provide an overview of expression patterns, making it simple to identify clusters or trends in gene expression across different experimental conditions or biological samples.

### 3.2. Protein-Protein Interaction (PPI) Network Construction and Module Analysis

To explore the physical and functional associations among DEG proteins, the STRING database was utilized. A minimum required interaction score of ≥0.4 was set, and the resultant interaction network was visualized using Cytoscape v3.8.2. The network comprised 264 nodes and 613 edges, with upregulated genes highlighted in red (cutoff logFC ≥ 1) and downregulated genes highlighted in green (cutoff logFC ≤ 1). Network analysis identified ATM, IL4, GATA3, and CTLA4 genes as highly interacted hub genes, with degrees of interaction ranging from 21 to 26.

### 3.3. Functional Enrichment Analysis using DAVID

Functional annotation of DEGs was performed using the DAVID v6.8 online server. KEGG pathway enrichment analysis and Gene Ontology (GO) classification were conducted to elucidate the biological roles of DEGs[14]. DEGs were found to be enriched in various biological processes, molecular functions, and cellular components. Specifically, upregulated genes were associated with processes such as steroid catabolism, immune response, and cytokine activity, while downregulated genes were implicated in pathways related to adaptive immune response and antigen binding[15]. Additionally, DEGs were found to be involved in diverse cellular components, including lysosomes, endosomes, and the endoplasmic reticulum. KEGG pathway analysis revealed enrichment in pathways such as steroid hormone biosynthesis, cytokine-cytokine receptor interaction, and JAK-STAT signaling pathway, among others, highlighting their potential roles in peanut allergen response[16, 17].

## 4. DISCUSSION

In this study, we delved into the differential gene expression between nineteen peanut allergens and five controls using the SRA ID SRX14128943 as a reference. Our analysis scrutinized a total of 46,694 differentially expressed genes (DEGs), from which we identified the top 444 significant DEGs for further investigation (Table 1). Notably, key genes such as ATM, IL4, GATA3, and CTLA4 emerged as highly interactive hub genes, suggesting their potential importance in the context of peanut allergy (Fig 4).

**Fig 4:**
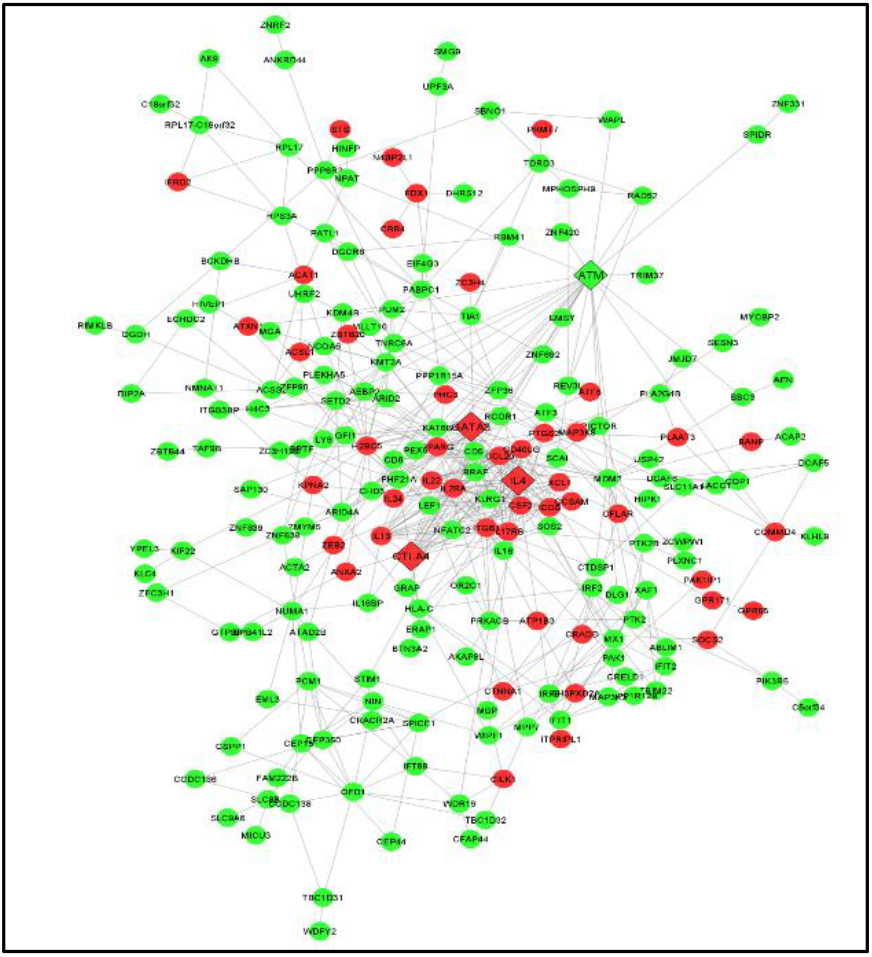
Gene network analysis is performed using String involves obtaining protein-protein interaction data to create a network graph in which genes are represented as nodes and interactions as edges. This network data is then able to be imported into Cytoscape for visualisation and analysis, allowing researchers to investigate gene interconnectivity, functional modules, and putative biomolecular mechanisms driving complex biological processes. Cytoscape provides strong tools for visualizing and analyzing biological networks, making it easier to understand gene connections and their implications in a variety of biological contexts.

**Fig 5:**
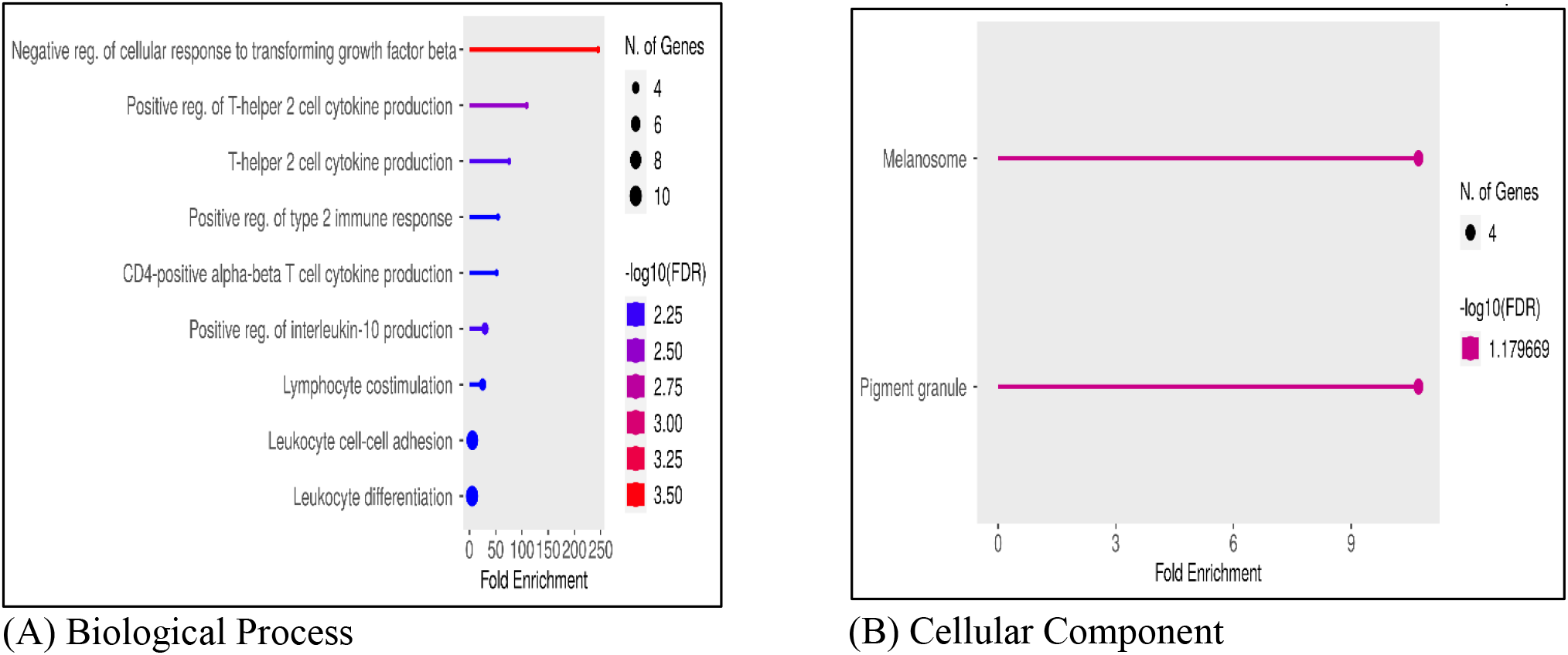
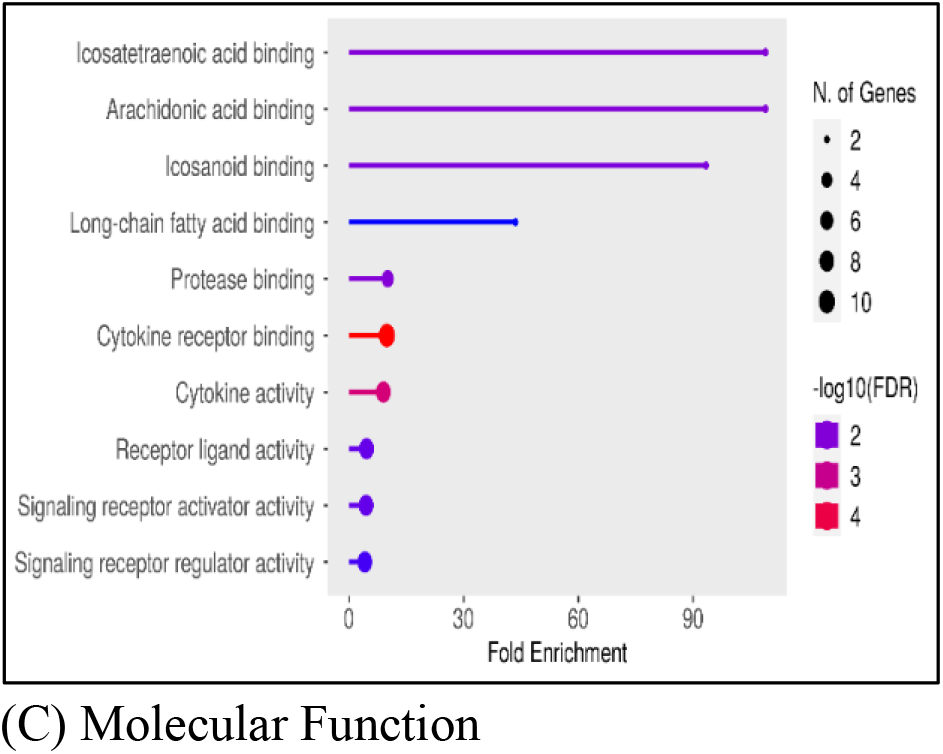
Shows the results of a functional enrichment study that focused on upregulated genes, highlighting the enriched (A)biological processes, (B) cellular components and (C) molecular functions associated with upregulated genes. The visualization provides insights into the biological pathways or mechanisms that may be activated or strengthened in response to experimental conditions, which aids in the understanding of gene expression changes.

**Fig 6:**
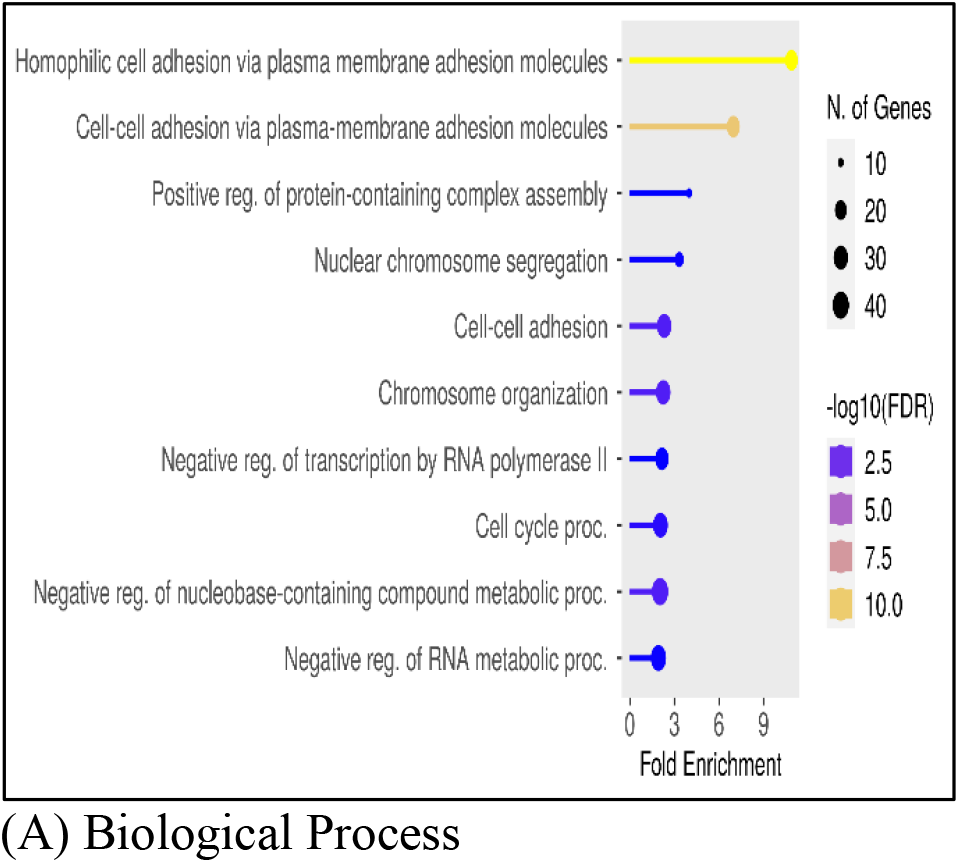
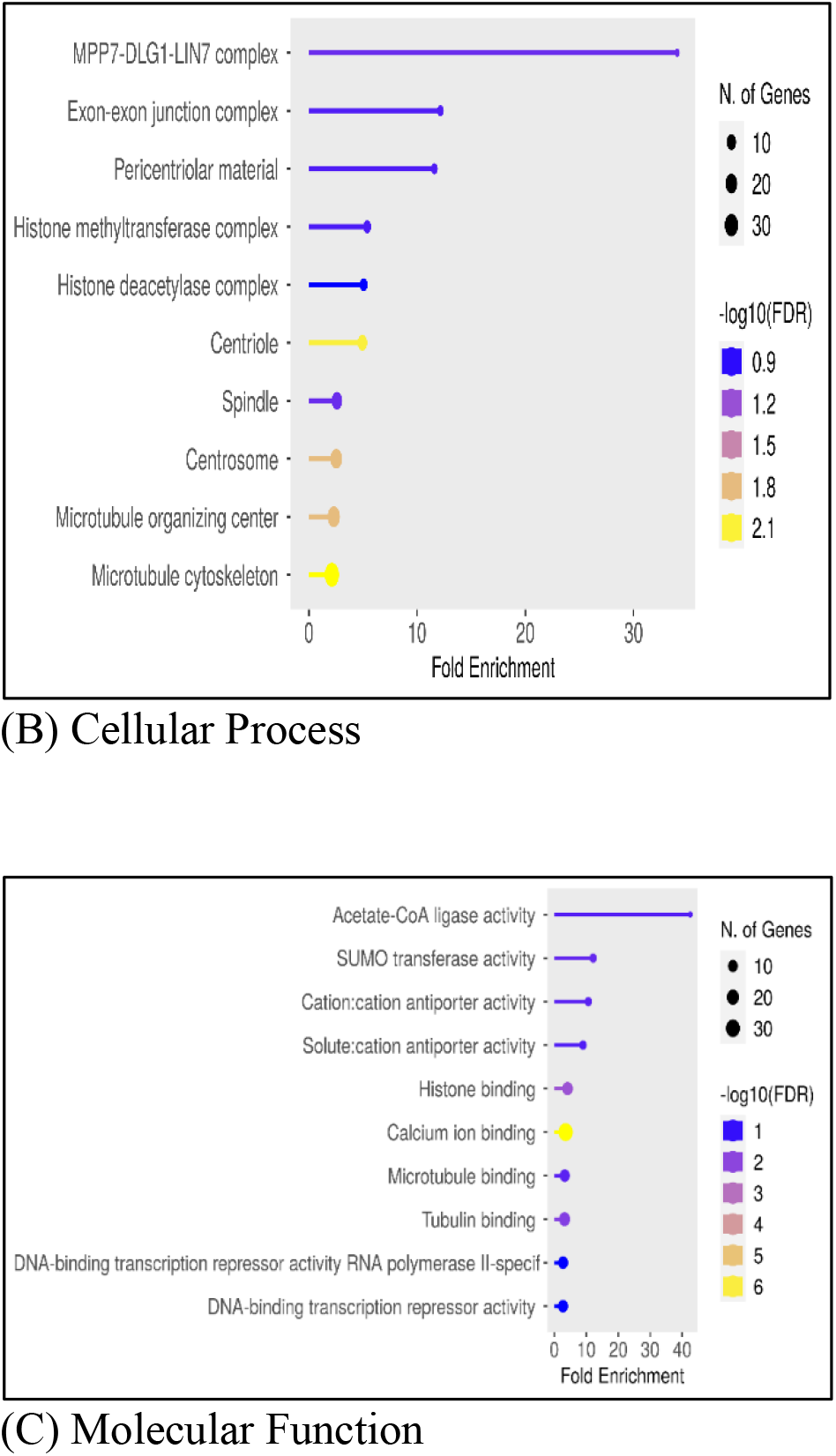
Shows the results of a functional enrichment study that focused on upregulated genes, highlighting the enriched (A)biological processes, (B) cellular components and (C) molecular functions associated with downregulated genes. The visualization provides insights into the biological pathways or mechanisms that may be activated or strengthened in response to experimental conditions, which aids in the understanding of gene expression changes.

**Fig 4:**
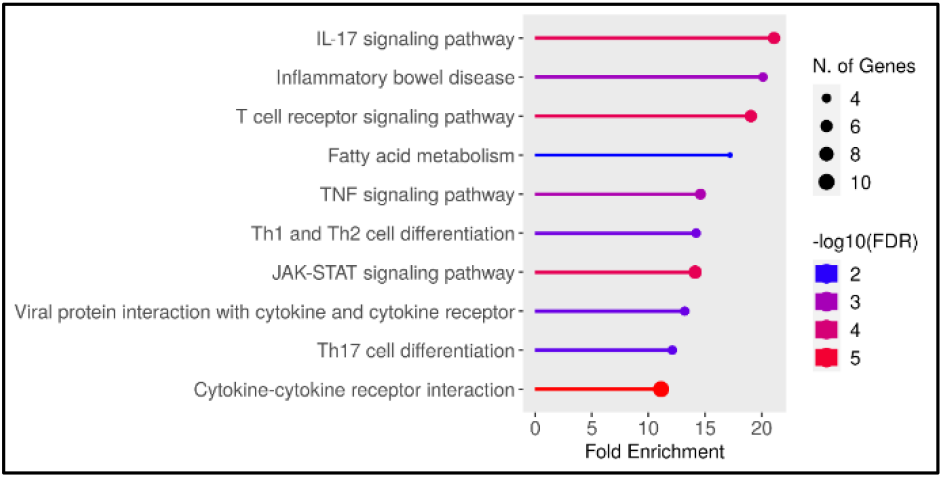
This figure depicts the results of a pathway enrichment analysis, emphasizing the enriched biological pathways linked to the experimental data. This graphic provides insights into the functional pathways that may be influenced or altered by the conditions under investigation.

Moreover, our examination revealed three seed nodes—IL4, IL13, CCL20, and CSF2— potentially involved in the regulation of pathways implicated in peanut allergy[18]. To elucidate the functional roles of these DEGs, we conducted Gene Ontology (GO) and Kyoto Encyclopedia of Genes and Genomes (KEGG) enrichment analyses, which highlighted their involvement in crucial pathways such as cytokine-cytokine receptor interaction, JAK-STAT signaling, and T cell receptor signaling pathways, among others[14, 17].

Focusing on specific genes such as STS, IL13, IL4, IL17RB, HLA-C, and AEN, we explored their roles in modulating immune responses and allergic reactions to food allergens. Steroid sulfatase (STS), for instance, implicated in steroid hormone metabolism, may contribute to allergic sensitization through immune modulation and epithelial barrier function. Similarly, cytokines IL-13 and IL-4 play pivotal roles in type 2 immune responses and allergic inflammation, with genetic variations in their genes associated with increased susceptibility to food allergies. IL17RB and HLA-C also contribute to allergic diseases through their roles in immune regulation and antigen presentation[19], while the AEN gene, involved in DNA repair and apoptosis regulation, may indirectly modulate allergic responses[18].

Further analysis of molecular functions (MF) and biological processes (BP) using GO revealed significant enrichment patterns among upregulated and downregulated DEGs. Upregulated genes were associated with cytokine receptor binding and leukocyte differentiation, while downregulated genes showed enrichment in calcium ion binding and negative regulation of nucleobase-containing compound metabolic processes, among others[20, 21]. Additionally, cellular component (CC) analysis identified enrichment in melanosome and microtubule cytoskeleton among upregulated and downregulated DEGs, respectively (Table 2 and Table 3).

**Table 2:**
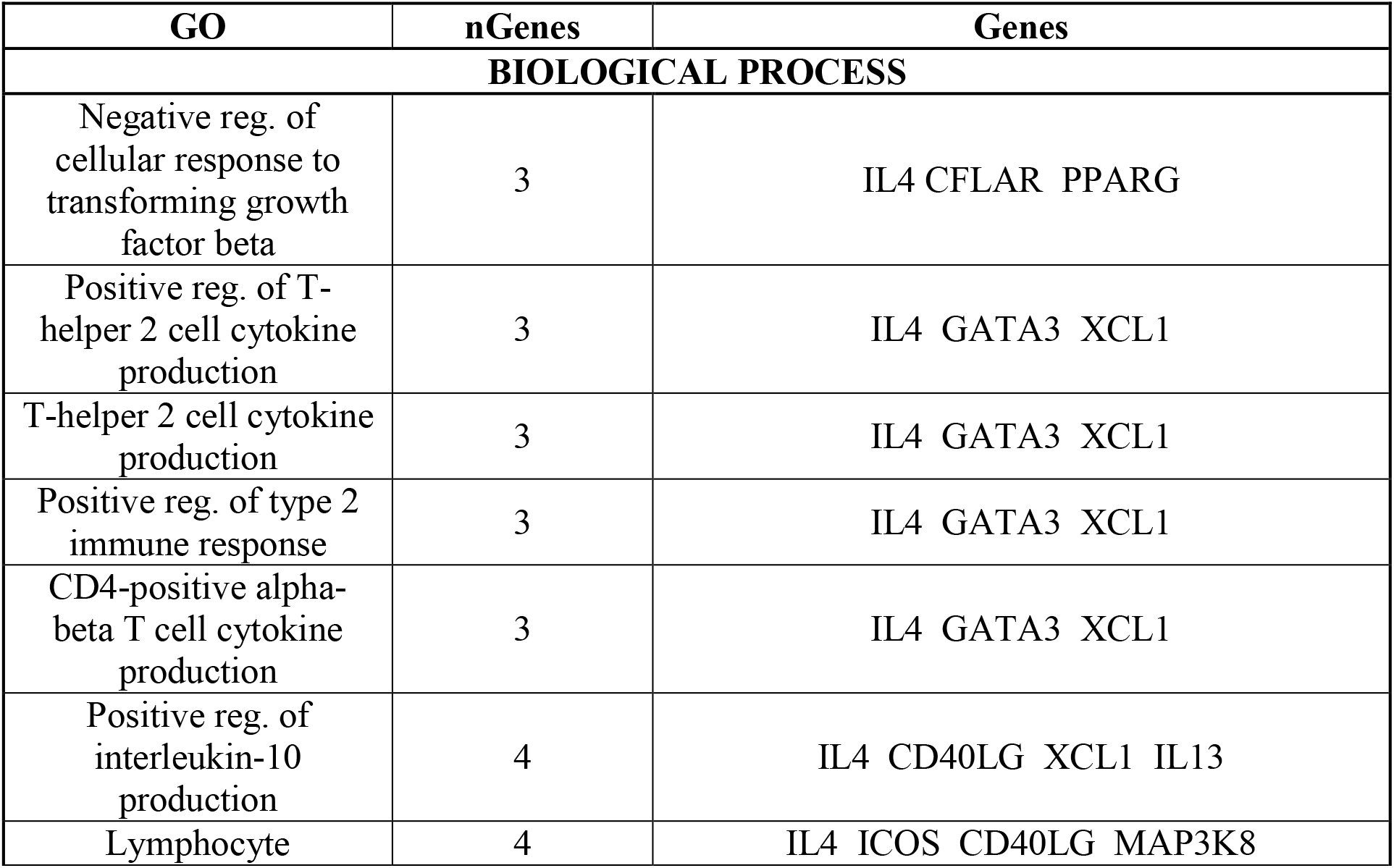
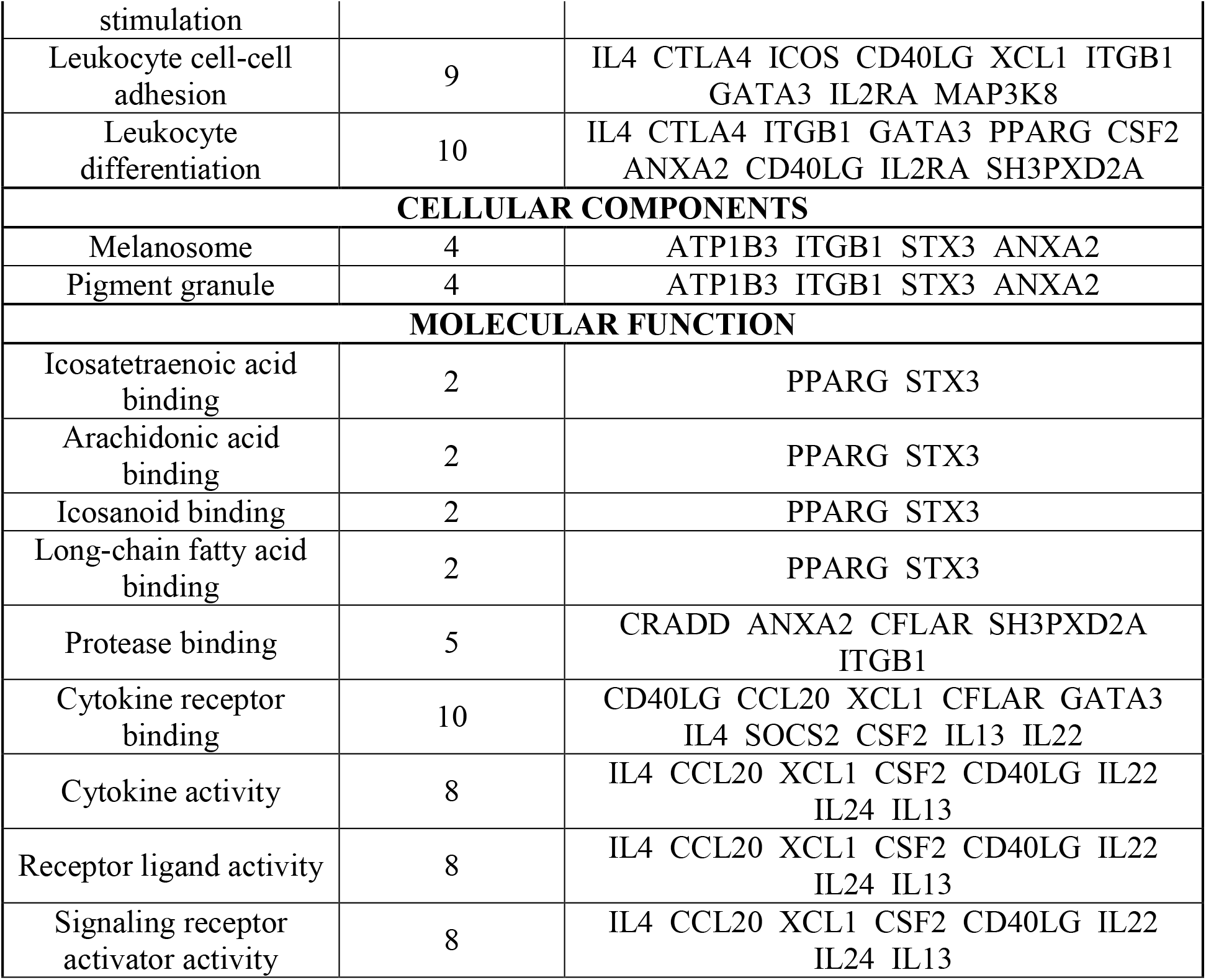
Enriched Gene Ontology (GO) Analysis Highlighting Upregulated Genes Identified via ShinyGO Database.

**Table 3:**
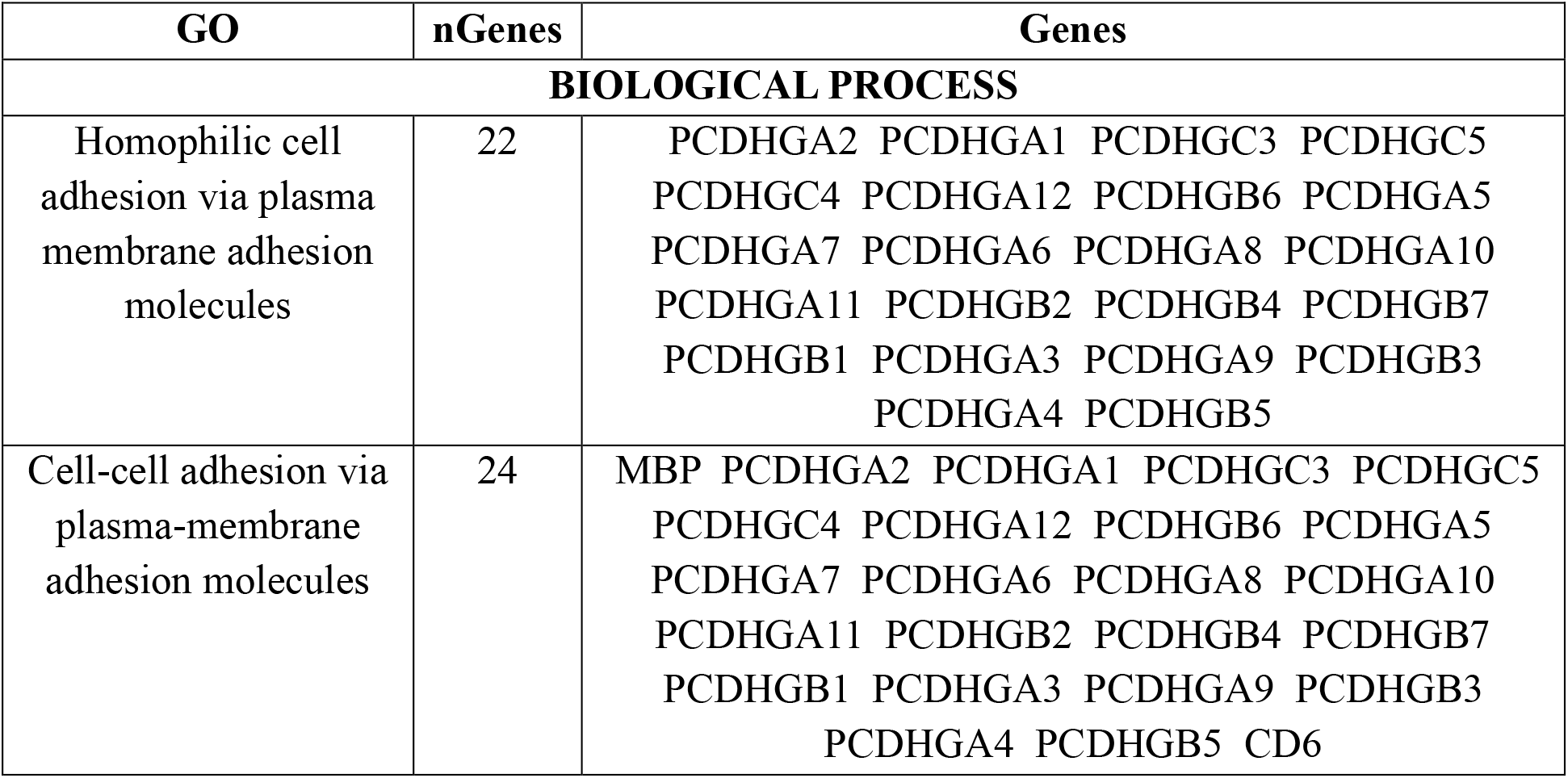
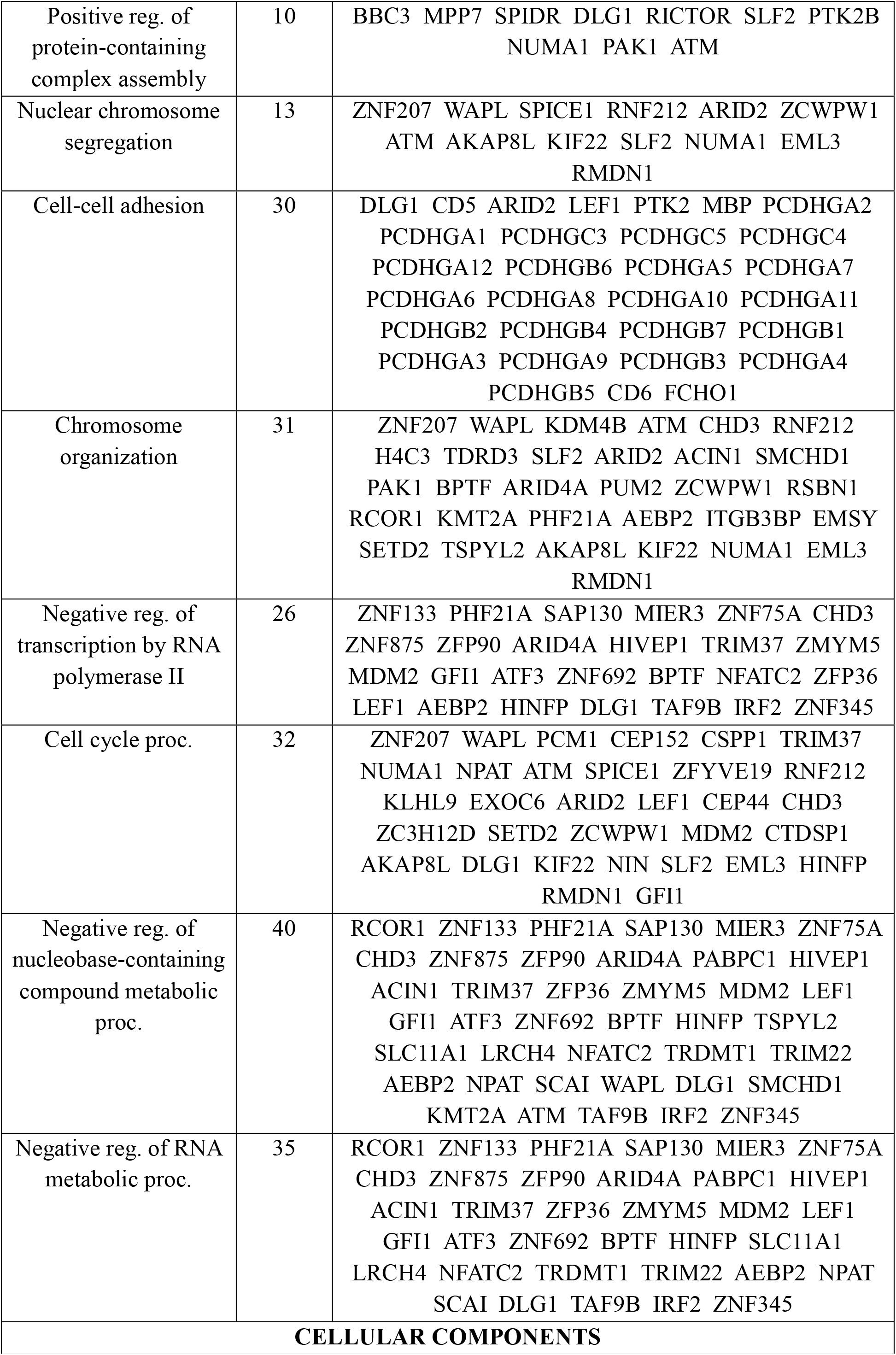
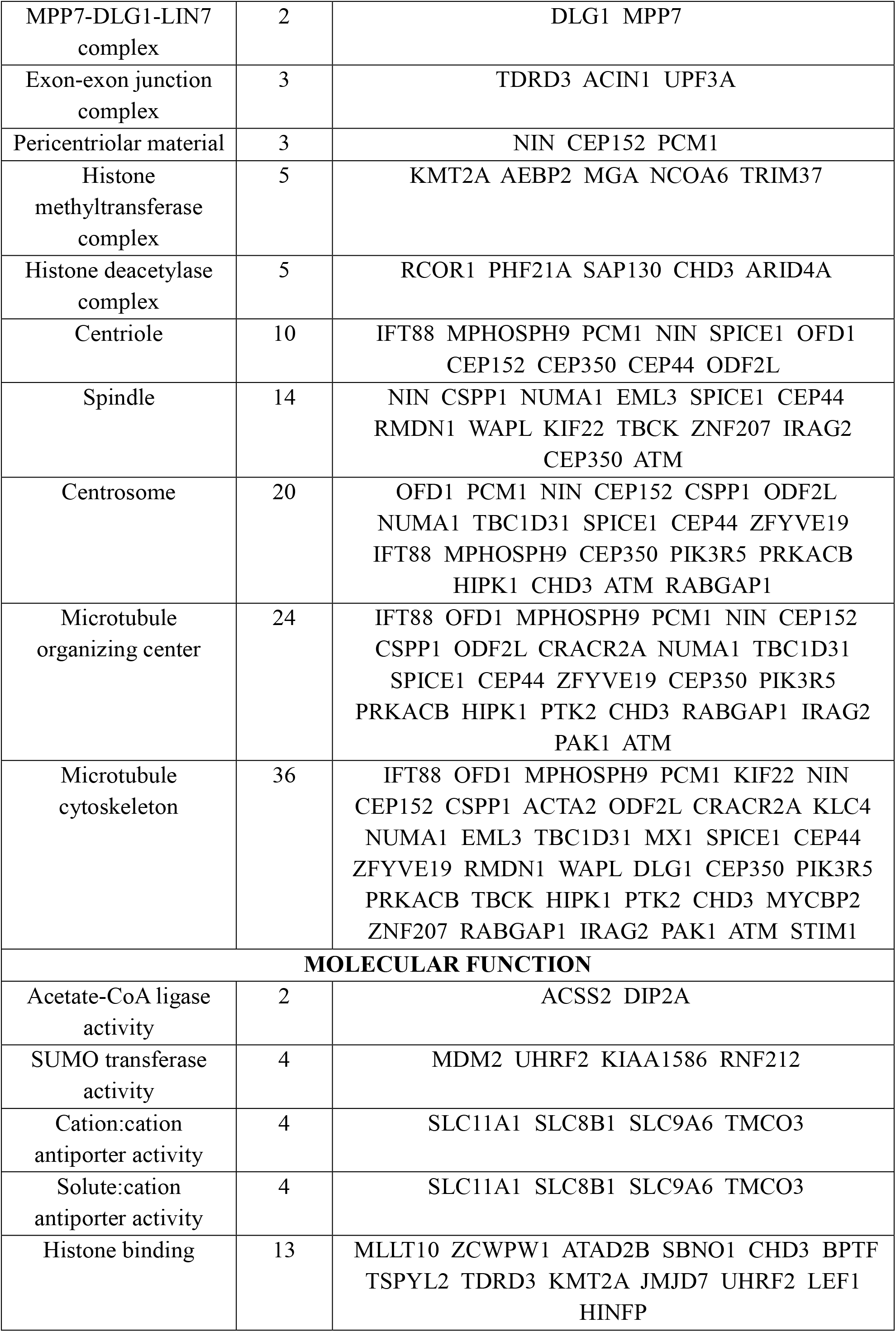
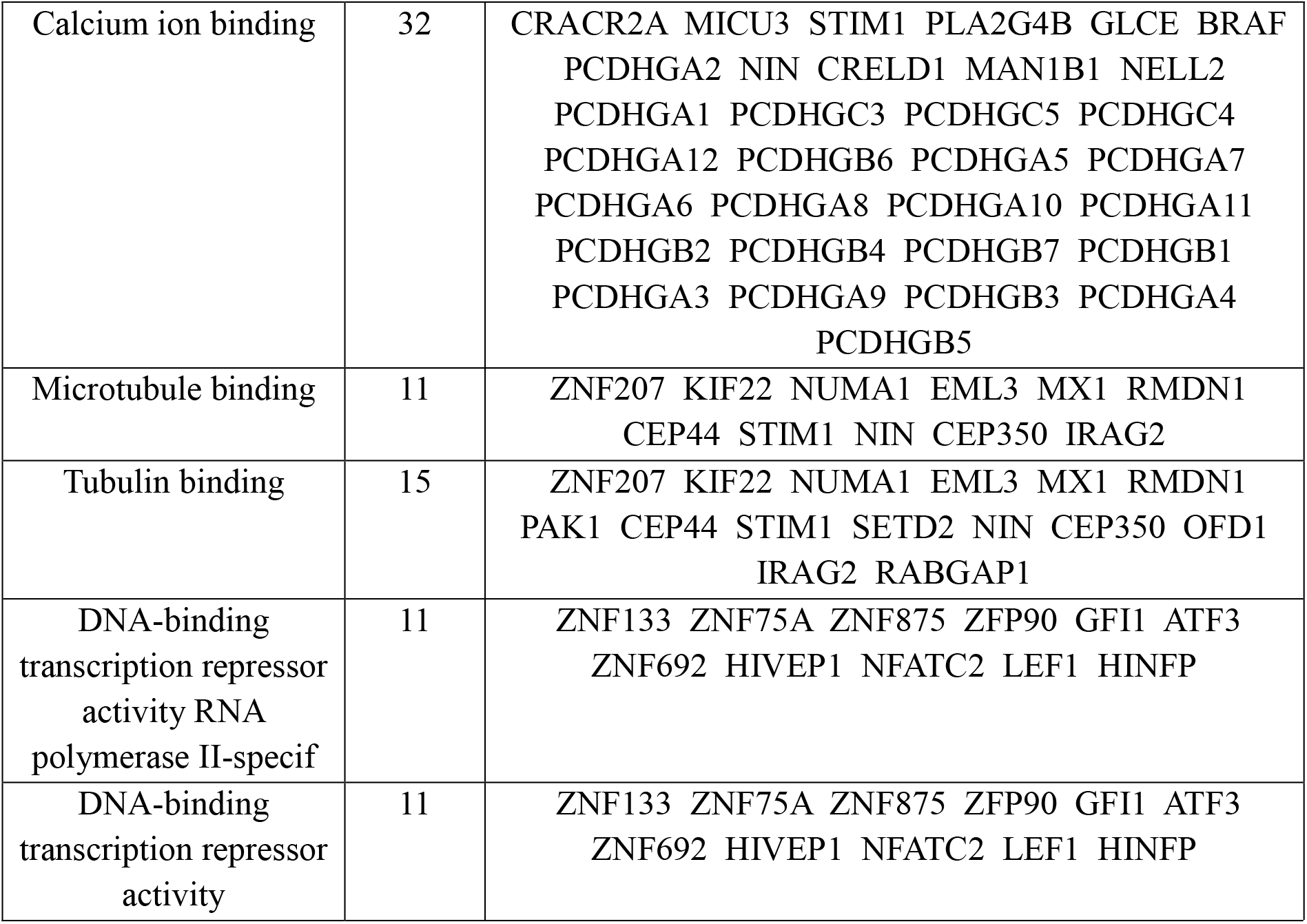
Highly enriched Gene Ontology(GO) Analysis for Downregulated genes from ShinyGO database.

**Table 4:** Pathway Enrichment Analysis Performed Using the ShinyGO Database.

| Pathways | nGenes | Genes |
| --- | --- | --- |
| hsa04657 IL-17 signalling pathway | 6 | CSF2 IL4 IL13 IL17RB PTGS2 CCL20 |
| hsa05321 Inflammatory bowel disease | 4 | GATA3 IL4 IL13 IL22 |
| hsa04660 T cell receptor signalling pathway | 6 | MAP3K8 CSF2 CTLA4 ICOS IL4 CD40LG |
| hsa01212 Fatty acid metabolism | 3 | ACSL1 ACAT1 CBR4 |
| hsa04668 TNF signalling pathway | 5 | MAP3K8 CSF2 PTGS2 CCL20 CFLAR |
| hsa04658 Th1 and Th2 cell differentiation | 4 | GATA3 IL2RA IL4 IL13 |
| hsa04630 JAK-STAT signalling pathway | 7 | IL24 CSF2 IL2RA IL4 IL13 IL22 SOCS2 |
| hsa04061 Viral protein interaction with cytokine and cytokine receptor | 4 | IL24 IL2RA CCL20 XCL1 |
| hsa04659 Th17 cell differentiation | 4 | GATA3 IL2RA IL4 IL22 |
| hsa04060 Cytokine-cytokine receptor interaction | 10 | IL24 CSF2 IL2RA IL4 IL13 IL22 IL17RB CCL20 XCL1 CD40LG |

This comprehensive exploration sheds light on the molecular mechanisms underlying peanut allergy and provides valuable insights for potential therapeutic targets and intervention strategies[1]. Further studies are warranted to validate these findings and translate them into clinical applications for improved management of food allergies[18].

## 5. CONCLUSION

In conclusion, our study offers a thorough examination of the molecular intricacies of peanut allergy, elucidating crucial differentially expressed genes (DEGs) and pathways linked to peanut-specific immune reactions. Through our systematic and bioinformatics-driven methodology, we have deepened our comprehension of the fundamental mechanisms driving allergic responses to peanuts. These findings present promising avenues for the creation of innovative therapeutic strategies and diagnostic biomarkers, which could significantly advance the management of peanut allergies.

## Data Availability

All data produced are available online at https://www.ncbi.nlm.nih.gov/sra/?term=SRX14128943

